# Seroprevalence of SARS-CoV-2 antibodies in broadcast media workers

**DOI:** 10.1101/2021.04.29.21256360

**Authors:** Paulo Ricardo Martins-Filho, Danilo Nobre da Silva, Danillo Menezes dos Santos, Márcia Santos Rezende, Jessica Paloma Rosa Silva, Josafá Bonifácio da Silva Neto, Dulce Marta Schimieguel, Lucindo José Quintans-Júnior, Jullyana de Souza Siqueira Quintans, Paula Santos Nunes, Adriano Antunes de Sousa Araújo

**Affiliations:** Investigative Pathology Laboratory, Federal University of Sergipe, Aracaju, Sergipe, Brazil; Health Sciences Graduate Program, Federal University of Sergipe, Aracaju, Sergipe, Brazil; Graduate Program in Pharmaceutical Sciences, Federal University of Sergipe, São Cristóvão, Sergipe, Brazil; Department of Pharmacy, Federal University of Sergipe, São Cristóvão, Sergipe, Brazil; Graduate Program in Applied Health Sciences, Federal University of Sergipe, Lagarto, Sergipe, Brazil; Journalism Sector, Radio UFS, Federal University of Sergipe, Aracaju, Sergipe, Brazil

**Author notes:** Address for correspondence: Prof. Paulo Ricardo Martins-Filho. Universidade Federal de Sergipe, Hospital Universitário, Laboratório de Patologia Investigativa. Rua Cláudio Batista, s/n. Sanatório. Aracaju, Sergipe, Brasil. CEP: 49060-100.

**Keywords:** COVID-19, SARS-CoV-2, Communications Media

## Abstract

In this cross-sectional study, we investigated the seroprevalence of SARS-CoV-2 antibodies in workers of radio and television (TV) in Sergipe state, Northeast Brazil. The study was conducted from December 1 to December 20, 2020, considered the beginning of the second wave of COVID-19 in the state. Our findings showed a high seroprevalence of SARS-CoV-2 antibodies in radio and TV workers, especially among those in the production and operation teams. Prevention and control protocols against COVID-19 should be revised and implemented by media companies. Broadcast media workers should be prioritized in COVID-19 vaccine strategies.

---

The COVID-19 pandemic has emerged as an unprecedented challenge for journalism activity.^1^ Apart from the uncertainty about job security, broadcast media workers are at increased risk of SARS-CoV-2 infection due to the daily routine of reporting in the field. Recently, the Press Emblem Campaign (PEC) – an international independent nonprofit and non-governmental organization -reported that more than 700 journalists died from COVID-19 in 72 countries since March 2020. Peru ranks first in the number of deaths (135), followed by Brazil (129) and Mexico (90) (https://www.pressemblem.ch/pec-news.shtml).

To the best of our knowledge, no seroepidemiological studies were conducted analyzing the presence of SARS-CoV-2 antibodies in broadcast media including workers of radio and television (TV). Surveillance of antibody seropositivity can allow to quantify the extent of infection in a population and proportion of people that remains susceptible to the virus. In this cross-sectional study, we investigated the seroprevalence of SARS-CoV-2 antibodies in radio and TV workers in Sergipe state, Northeast Brazil. The study was conducted from December 1 to December 20, 2020, considered the beginning of the second wave of COVID-19 in the state.

After obtaining written informed consent to participate, individuals were interviewed using a structured questionnaire that included demographic and clinical features. Then, venous blood was collected aseptically using venipuncture and a fluorescence immunoassay (FIA) (iChroma II, BioSys + Kovalent) for qualitative detection and differentiation of IgM and IgG antibodies against SARS-CoV-2 was performed. A result was considered negative if the automated reader obtained a readout <0.8, indeterminate if ≥0.8 and <1.1, and positive if ≥1.1. The sensitivity and specificity of the FIA are 95.8% and 97.0%, respectively, according to the manufacturer when assessed on 46 SARS-CoV-2 positive patients and 131 negative controls (http://www.biosys.com.br/wp-content/themes/transport/covid-19-images/SOLUCAOPOCT-COMPLETA-COVID-19.pdf).

The main outcome in the present study was seroprevalence expressed as the proportion of individuals who had a positive result in the FIA. Broadcast media workers were grouped according to the occupation activity as following: (1) production team (PT); (2) reporting team (RT); and operation team (OT). Pearson’s chi-square test was performed to examine differences in seroprevalence by type of media work. Significance level was set at 5%. Analysis was performed by using R software (version 3.5.3; R Foundation for Statistical Computing, Vienna, Austria). This study was approved by the Ethics Committee of the Federal University of Sergipe (protocol number 33095120.4.0000.5546).

The study included a convenience sample of 113 broadcast media workers (62 PT, 22 RT, and 29 OT). The median age (interquartile range [IQR]) was 39 years (IQR, 30.0 – 48.0) and most of them were male (n = 79, 70.0%). The presence of comorbidities (diabetes, hypertension, cardiac disease, or other chronic condition) was reported by 29 (25.7%) individuals. Forty-two (37.2%) workers described close contact with people presenting SARS-CoV-2 infection in the last two weeks of the serological assay and the use of facial mask during work activities was reported in most cases (n = 110, 97.3%).

Twenty-eight broadcast media workers had detectable levels of SARS-CoV-2 antibodies (11 IgM+, 6 IgM+/ IgG+, and 11 IgG+) and the estimated seroprevalence was 24.7% (95% CI 17.7 – 33.5). There was no statistical difference in the seroprevalence between occupational activities (p = 0.715), but OT (27.6%, 95% CI 14.7 – 45.7) and PT (25.8%, 95% CI 16.6 – 37.9) presented higher estimates than RT (18.2%, 95% CI 7.3 – 38.5) (Table 1).

**Table 1.** Clinical characteristics and seroprevalence of SARS-CoV-2 antibodies among broadcast media workers.

| Variable | All media workers<br>(n = 113) | Production team<br>(n = 62) | Reporting team<br>(n = 22) | Operation team<br>(n = 29) |
| --- | --- | --- | --- | --- |
| Age (median, IQR) | 39.0 (30.0 – 48.0) | 40.0 (33.3 – 47.0) | 42.0 (35.8 – 51.8) | 30.0 (25.0 – 39.0) |
| Sex (male) | 79 (70.0%) | 40 (64.5%) | 17 (77.3%) | 22 (75.9%) |
| Comorbidities | 29 (25.7%) | 19 (30.7%) | 6 (27.3%) | 4 (13.8%) |
| Close contact with someone who had COVID-19 | 42 (37.2%) | 29 (46.8%) | 6 (27.3%) | 7 (24.1%) |
| Use of face mask | 110 (97.3%) | 60 (96.8%) | 21 (95.5%) | 29 (100.0%) |
| Serological analysis |  |  |  |  |
| IgM+ | 11 (9.7%) | 7 (11.3%) | 2 (9.1%) | 2 (6.9%) |
| IgM+/IgG+ | 6 (5.3%) | 4 (6.5%) | 0 | 2 (6.9%) |
| IgG+ | 11 (9.7%) | 5 (8.1%) | 2 (9.1%) | 4 (13.8%) |
| Seroprevalence (CI 95%) | 24.7 (17.7 – 33.5) | 25.8 (16.6 – 37.9) | 18.2 (7.3 – 37.9) | 27.6 (14.7 – 45.7) |

Despite the massive interest in social networking services for information sharing and Internet use, traditional media remains the most important channel through which COVID-19 information is communicated.^2^ Faced with the need for real-time coverage of COVID-19 information in several settings, broadcast media workers are at increased risk of SARS-CoV-2 infection. Ensuring that a media worker has the personal protective equipment (PPE) and support they need to work safely has become critical during this unprecedented global sanitary crisis.

In this study, we found a high seroprevalence estimate in radio and TV workers and a large proportion of individuals with serological results suggestive of active phase or recent SARS-CoV-2 infection. These findings indicate a high exposure of broadcast media workers to the SARS-CoV-2 and the circulation of these individuals in the work environment unaware that they are infected with the virus. It has been shown that SARS-CoV-2 may spread asymptomatically in a population even under social distancing restrictions.^3^

Interesting, our results showed a lower seroprevalence for SARS-CoV-2 antibodies among journalists, reporters, and videographers of broadcast media. It is possible that RT, considered the high-risk group for SARS-CoV-2 infection, is more careful regarding COVID-19 protective measures due to the need for contact with the external public during work activities. In contrast, PT and OT usually work inside TV newsrooms which may increase the risk of infection in an indoor environment. There is evidence that closed indoor spaces with minimal ventilation rate provide an ideal environmental for SARS-CoV-2 transmission.^4,5^

This study showed a high seroprevalence of SARS-CoV-2 antibodies in radio and TV workers, especially among those in the production and operation teams. Prevention and control protocols against COVID-19 should be revised and implemented by media companies. Broadcast media workers should be prioritized in COVID-19 vaccine strategies. Further seroepidemiological studies should evaluate the exposure of freelancers and print and digital media professionals to SARS-CoV-2 infection.

## Data Availability

The data that support the findings of this study are available on request from the corresponding author.

## Acknowledgements

To all frontline workers who are facing the COVID-19 pandemic. This study is part of the EpiSERGIPE project which is supported by grant SES/FAPESE/UFS 001/2020. The funding sources had no role in the design and conduct of the study; collection, management, analysis, and interpretation of the data; preparation, review, or approval of the manuscript; and the decision to submit the manuscript for publication.

